# Analyzing Healthcare Safety Climate Through A Collaborative Filtering of Safety Attitudes Questionnaire for A Hospital Case Study

**DOI:** 10.1101/2021.02.11.20202440

**Authors:** Huiyu Huang, Chun-An Chou, Shao-Jen Weng, Chieh-Liang Wu

## Abstract

**Background:** Patient safety climate in healthcare is an important element to improve quality of current healthcare system globally. Safety Attitudes Questionnaire (SAQ), one of measurable tools for patient safety climate, is applied as an effective research survey to collect opinions and information from medical workers in a long term.

**Method:** SAQ data in this study consists of medical employee scores to 46 questions in 8 aspects or dimensions and personal information during 2014 to 2017 from a teaching hospital. A quantitative analysis with a core user based collaborative filtering method is conducted to analyze the SAQ data for discovering the latent inter-related pattern among different SAQ aspects of sub-sectors in this hospital for four years.

**Results:** There are not interrelationships between these 8 independent aspects for wards at different level (A, B and C) consistently across 4 years. The correlation patterns of aspects are not strong consistent over years for wards.

**Conclusion:** Observations in this study can be used to support understanding patient safety climate assessment. They are contributed to further study in this area and understand SAQ aspects dynamically in a real hospital case.

## INTRODUCTION

Numerous volumes of errors and accidents caused by unsafe medical service and their serious negative effects across the world [1–5] are reminding people to focus on patient safety culture or climate because of urgency and necessity to eliminate foreseen medical errors and incidents for seeking a satisfactory patient safety standard and environment. In other words, ignorance of these medical issues causes a poor reputation and investing plenty of additional capitals for remedy, which is not regarding as an advisable management strategy anymore. Patient safety culture is believed to be a relatively stable environment affecting staff to meet certain working standards and optimizing working procedures for provision of health services.

### Safety Measurement - Safety Attitudes Questionnaire (SAQ)

In the past 20 years, safety attitudes questionnaire (SAQ) is one of the most universal instruments to evaluate patient safety culture with regarding of psychometric properties from medical employees’ opinions [6]. SAQ was originally designed by Sexton et al. in 2006, eliciting caregiver attitudes through the six factor analytically derived climate scales: teamwork climate, safety climate, job satisfaction, perceptions of management, working conditions, and stress recognition with 60 items and demographics information (e.g., age, sex, experience, and nationality) with numbered or open-end answers as a benchmark in psychometric surveys to measure these attitudes [7]. It is a useful management tool to measure the safety climate timely from medical workers in a medical place through a relatively convenient and fast mode. Many versions have been developed to explore and improve healthcare quality in different medical organizations and its results assist the continuous adjustment of management strategies [8,9]. SAQ has been translated into multiple languages such as Greek-Cypriot [10], Norwegian [11], Swedish [12], Turkish [13], Indian [14], Chinese [15] etc. and adjusted to diversified setting in medical places such as SAQ-A [16], SAQ -ICU [10,17], cross-sectional or national investigation, long-term survey, etc. These SAQs in different versions have been tested and proved their validity and reliability generally via Cronbach’s alpha in many medical places in different countries [14,15,18–23]. Apparently, it is a global trend for improving safety quality and selecting SAQ to measure conditions and analyze the situation in this territory.

This study adopts a Chinese version of the SAQ to evaluate the patient safety culture. This Chinese version of the SAQ was developed firstly by the Joint Commission of Taiwan in 2008, with 6 dimensions and 30 questions based on the short form of the SAQ to assess the patient safety culture in healthcare organizations on a yearly basis [24–26]. The Chinese version of the SAQ has been the official questionnaire used for assessing the patient safety culture in Taiwan since 2008 [26]. In 2014, three hospital-level aspects of safety culture from the Agency for Healthcare Research and Quality, which were originally included in the previous version, have been removed. Two dimensions, including the emotional exhaustion and work–life balance, have been added [24].

### Analysis of SAQ Data

To the best of our knowledge, there are basically two types of analysis methods: statistical methods and other methods. The statistical methods such as descriptive statistics including mean, medium, standard deviation, and frequency [14,15], linear regression[24], confidential intervals, analysis of variance (ANOVA), Student’s t-test [23], Pearson correlation [14,15], and exploratory factor analysis are usually applied on SAQ data to support some important conclusions. Other methods are ad hoc analysis, decision-making trial and evaluation laboratory. In this study, we propose a new quantitative analysis method based on the core concept of User-based Collaborative Filtering (UB-CF). UB-CF has been a widely adapted recommender system relying on the rating score (e.g. 1 is the lowest recommendation/satisfaction and 5 is highest recommendation/ satisfaction) defined firstly as an independent area since the mid-1990s[27]. The three assumptions of Collaborative Filtering algorithms are: (1) ones have similar preferences and interests; (2) their preferences and interests are stable; and (3) their choice can be estimated according to their past preferences [28].

Many conclusions are deduced after the SAQ analysis is implemented. Gallego et al. identified that elderly staff have lower stress recognition scores, indicating they did not think stressors would have significant effects on personal performance [29]. Kirwan, Matthews, and Scott stated that nurses with different education levels have different patient outcomes [30]. Furthermore, Hamdan proposed that gender and age are critical factors impacting patient safety [31]. Wu et al. reported that gender, age, job position, job status, and education are critical factors influencing the patient safety culture [32]. Lee et al. believed that medical staff with different demographic variables might have different perceptions on patient safety culture and using a longitudinal study to assess patient safety culture enables hospital management to trace the performance and trends on a timely basis [33].

In this study, we three-fold objectives are to provide an analysis method from a more date mining angle for mimicking inner patterns of SAQ data, identify interrelationships between aspects or aspects and overall evaluated levels (i.e. A, B and C) in terms of staff’s evaluation in specific wards of a partner teaching hospital, to explore some significative standards and results to support aspect selection for improving SAQ analysis in a real hospital case study.

## METHODS

### Data Acquisition

The survey data is collected from the quarterly evaluation of medical care teams or sub-sectors from a partner teaching hospital in Taiwan and is presented annually from 2014 to 2017. For privacy protection, the study was approved by the Institutional Review Board (IRB) of Taichung Veterans General Hospital (IRB TCVGH No. CW17045A). The respondents are registered nurses and medical administrators in this hospital and are changing due to employee turnover in these four years. The statistics of these respondents from selected wards (i.e. W1-W8) are presented in **Table 1** and **Tables 1-8** in the supplementary file. Most respondents are registered nurses and the rest of them are medical administrators in these wards.

**Table 1.** The number of medical staffs in these wards in the four years

|  | 2014 | 2015 | 2016 | 2017 |
| --- | --- | --- | --- | --- |
| W1 | 19 | 23 | 18 | 14 |
| W2 | 20 | 23 | 22 | 22 |
| W3 | 34 | 41 | 39 | 34 |
| W4 | 20 | 21 | 21 | 18 |
| W5 | 37 | 39 | 42 | 46 |
| W6 | 31 | 34 | 32 | 29 |
| W7 | 26 | 27 | 26 | 28 |
| W8 | 31 | 30 | 31 | 22 |

### Measure

SAQ applied in this paper collects demographic information and opinions about patient safety issues, medical errors, and event reports from medical workers. The content of SAQ designed for achieving this goal consists of 46 questions in 8 aspects (i.e., dimensions) and some other questions related to personal information such as gender, age, job status, job position, experience in organization, and experience in position. Concise definitions for Teamwork Climate (TC), Safety Climate (SC), Job Satisfaction (JS), Stress, Recognition (SR), Emotional Exhaustion (EE), Perception of Management (PM), Working Conditions (WC), Work and Life Balance (WB) aspects are illustrated in **Table 2** and the specific questions in them are showed in **Table 9** in the supplementary file. The marking system for all questions in first seven dimensions are five-point Likert scale ranging from strongly agree (5), agree (4), neutral (3), disagree (2), strongly disagree (1) with 0 as not applicable and for other questions in the last aspect are frequency such as never (4), rarely (3), sometimes (2), most of the time (1) with 0 as not applicable.

**Table 2.**
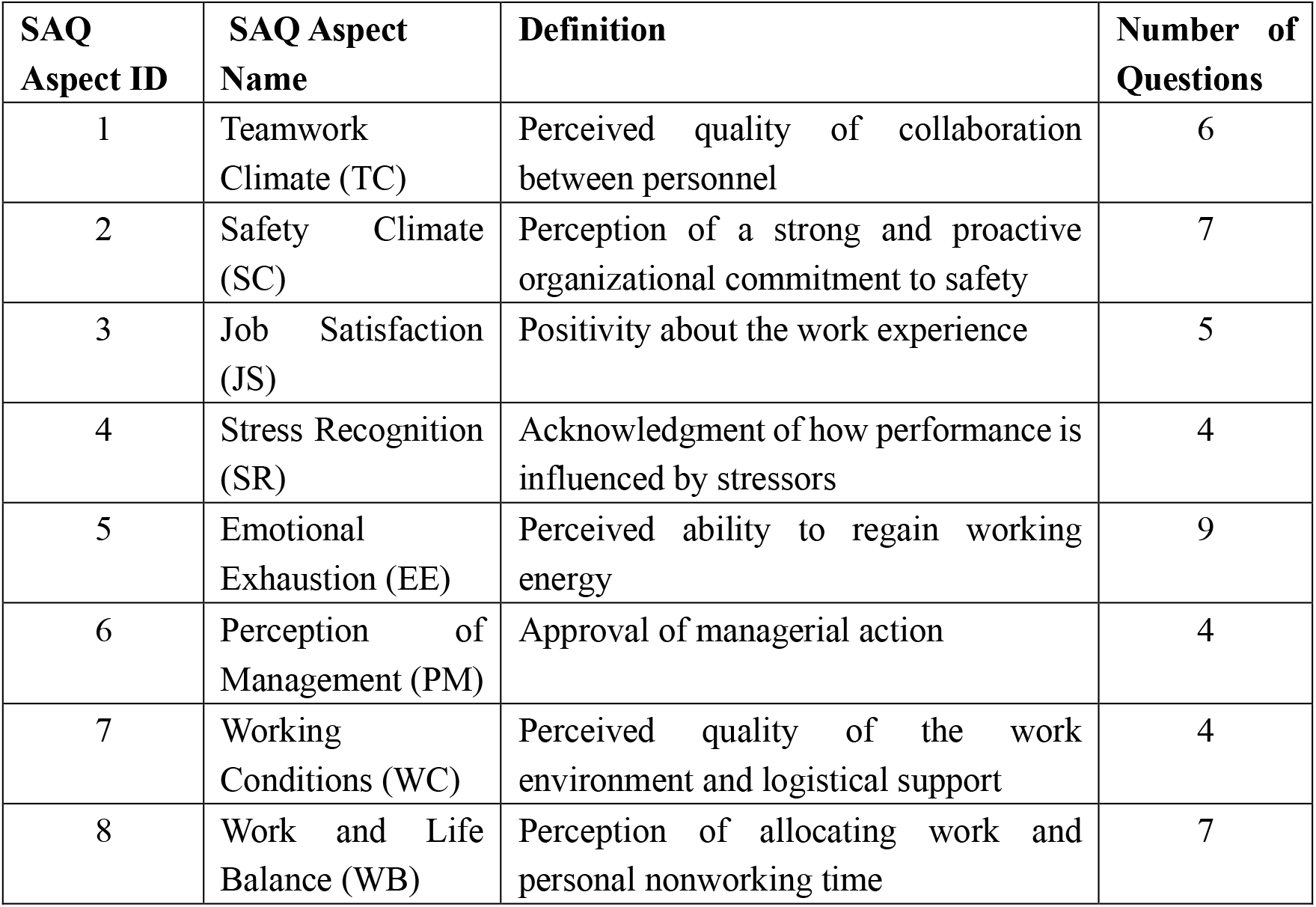
The introduction of aspects in the SAQ [18]

### Analysis

#### User based collaborative filtering (UB-CF) based Analysis

User based collaborative filtering (UB-CF) is a common and helpful method widely applied in recommendation systems because it contributes to search other users who have similarity in taste with a certain user and then introduce items that a certain user interested in these similar users. This method is centered around a statistical similarity measure to search the nearest neighbors of the object user and then basing on the item rating rated by the nearest neighbors to estimate the unknown item rating rated by the object user. It recommends new items based on users’ past behaviors and contains memory. It has demonstrated some successes in real-life applications integrated in other machine learning techniques such as the famous Amazon recommender system [34].

To conduct UB-CF based analysis, the similarity for each responder is required firstly. There are actually several measures to calculate the similarity, but only the cosine similarity is used for the following computation. The cosine similarity measure is acquiring angles between two vectors of ratings and a smaller angle is regarded as a greater similarity[35]. Its formula is

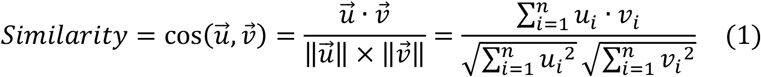

where 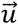 and 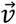 are two vectors containing the rates to *n* (i.e., 46) questions excluding questions related to personal information in SAQ given by two different respondents respectively and *u*_*i*_ or *v*_*i*_ is a score the respondent *u* or *v* given to question *i*. Then top-n-best neighbors served as recommenders are elected based on similarity measure and an assumed number *k* (i.e. the square root of the size of each assigned training set) [35]. The estimation result of a UB-CF model is computed using the following equation:

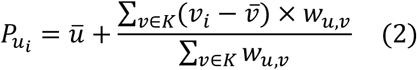

for a rate given to each question from a respondent denoted by *u*. Here *K* is a set containing *k* most similar respondents compared with rates given by respondent *u* and *w*_*u,v*_ is the similarity between the respondent *u* and *v*. In addition, the mean value of rates for 46 questions evaluated by the respondent *u* or *v* is referred as 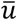 or 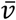 in Equation (2), respectively.

The performance measures for a UBCF model are mean absolute error (MAE), accuracy, specificity and sensitivity. The computation method for the MAE is

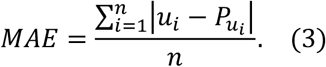

Other three measures are required to divide rates into some categories in advance. In this study, the rating ranges 4-5, 3-4, 1-3 are classified into three groups as positive attitude, neutral attitude and negative attitude correspondently. The accuracy is obtained in a percentage number representing the ratio of the number of estimated rates accurately in the total number of estimated rates for three groups. The specificity is a proportion of the number of estimated rates correctly in the total number of estimated rates only within the negative attitude group and the sensitivity is a similar proportion but within the positive attitude group.

#### General Procedure

The percentage value of each aspect for each ward is calculated based on the following equation:

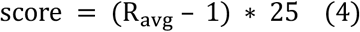

where *R*_*avg*_ is mean value of questions in each aspect for each sub-sector in general wards in 2014-2017 years. A percentage value of all aspects for each ward is the average of all percentage values of all aspects for this ward in each year and is able to be classified into three levels: A, B, and C. The classification criterion is that the level A contains scores above 88.5 (Excellent), scores between 80 and 88 are belong the level B (Good) and other scores are categorized into the level C (Fair). The trends of levels for several wards in Table 3 are contributed to complete some comparisons in further studies. For example, W1 is the only one that is always in level A in four years. W5 is always in the level B in four years. W3 is in the level B in 2014, but it is the level A in 2015 and then it goes back to level B in the following two years. W7 goes from the level C to the level A in the first three years and then back to the level B in 2017. W6 stays in the level B in three years and W8 is also in the level C in three years.

**Table 3.** The scores and levels for medical teams in general wards in four years

|  | 2014 | 2015 | 2016 | 2017 |
| --- | --- | --- | --- | --- |
| W1 | 89.02(A) | 88.50(A) | 98.25(A) | 98.33(A) |
| W2 | 93.04(A) | 88.25(B) | 73.42(C) | 82.00(B) |
| W3 | 82.53(B) | 89.25(A) | 85.58(B) | 86.58(B) |
| W4 | 80.50(B) | 83.33(B) | 88.75(A) | 88.92(A) |
| W5 | 86.41(B) | 86.42(B) | 86.08(B) | 86.08(B) |
| W6 | 84.25(B) | 79.00(B) | 76.83(C) | 79.29(B) |
| W7 | 76.07(C) | 81.42(B) | 91.08(A) | 87.08(B) |
| W8 | 82.71(B) | 77.42(C) | 68.83(C) | 67.42(C) |

These eight wards in **Table 3** are selected as subsets to be analyzed based on the types of sub-vectors in general wards from the dataset in every year. Then UB-CF model is implemented one hundred times to estimate scores representing answers to questions in the remaining seven aspects in term of scores representing answers to questions in a selected aspect for respondents in a testing set of that subset by inputting 80% of each subset selected randomly as a training set and the rest of it as a testing set in each year. To assess the estimation performance, mean absolute error (MAE), accuracy, specificity (<=3) and sensitivity (>=4) are perfect indicators for analyzing each question. The best associations between 8 aspects are derived by the smallest average values of MAEs and the largest average values of accuracies after the output (i.e. the average values of MAEs, accuracies for each questions) of the model is filtered by using a specific range. In addition, the range of mean values of MAEs is from 0 to 4 and the range of mean values of accuracies is from 0 to 1 since 0s as not applicable are considered as missing values in this analysis. The analytic framework is displayed in **Figure 1**.

**Figure 1.**
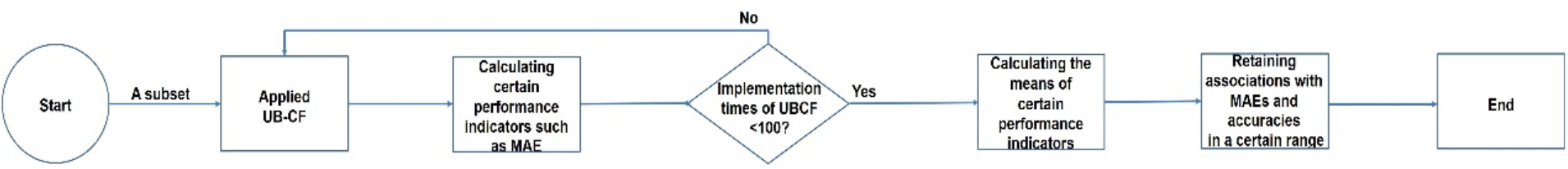
The proposed analytic framework.

## RESULTS

The mean values of MAEs and means of accuracies are selected for every aspect in the four years only when the means of MAEs are smaller than 1 and the means of accuracies are larger than 0.7. In this range, the numbers of existing associations between each two aspects in these eight aspects are collected for inferring the degree of importance of each aspect in the whole system and determining a role of each aspect. For each ward in these eight wards, these metrics are recorded into four tables representing relationships of aspects from 2014 to 2017. In every table, the aspect numbers in the first column are effects receivers of other aspects and aspect numbers in the first row are causes of other aspects. The names of aspect 1 to 8 are Team Climate, Safety Climate, Job Satisfaction, Stress Recognition, Emotional Exhaustion, Perception of Management, Working Conditions, Work-life Balance respectively. Heatmaps in **Figures 2-9** corresponding to these tables are generated to identify characters of the selected wards. The significance of each aspect is calculated by adding the amount of associations belong to each aspect together including the associations estimated by it and its associations estimated by other aspects. The role of each aspect is decided by positive or negative value obtained by using the number of associations estimated by it to subtract the number of its associations estimated by other seven aspects. If the outcome value is positive, the aspect is the cause to other aspects in the system. Otherwise, the aspect is a receiver of other aspects in this system. These results are showed in **Table 10** in the supplementary file.

**Figure 2.**
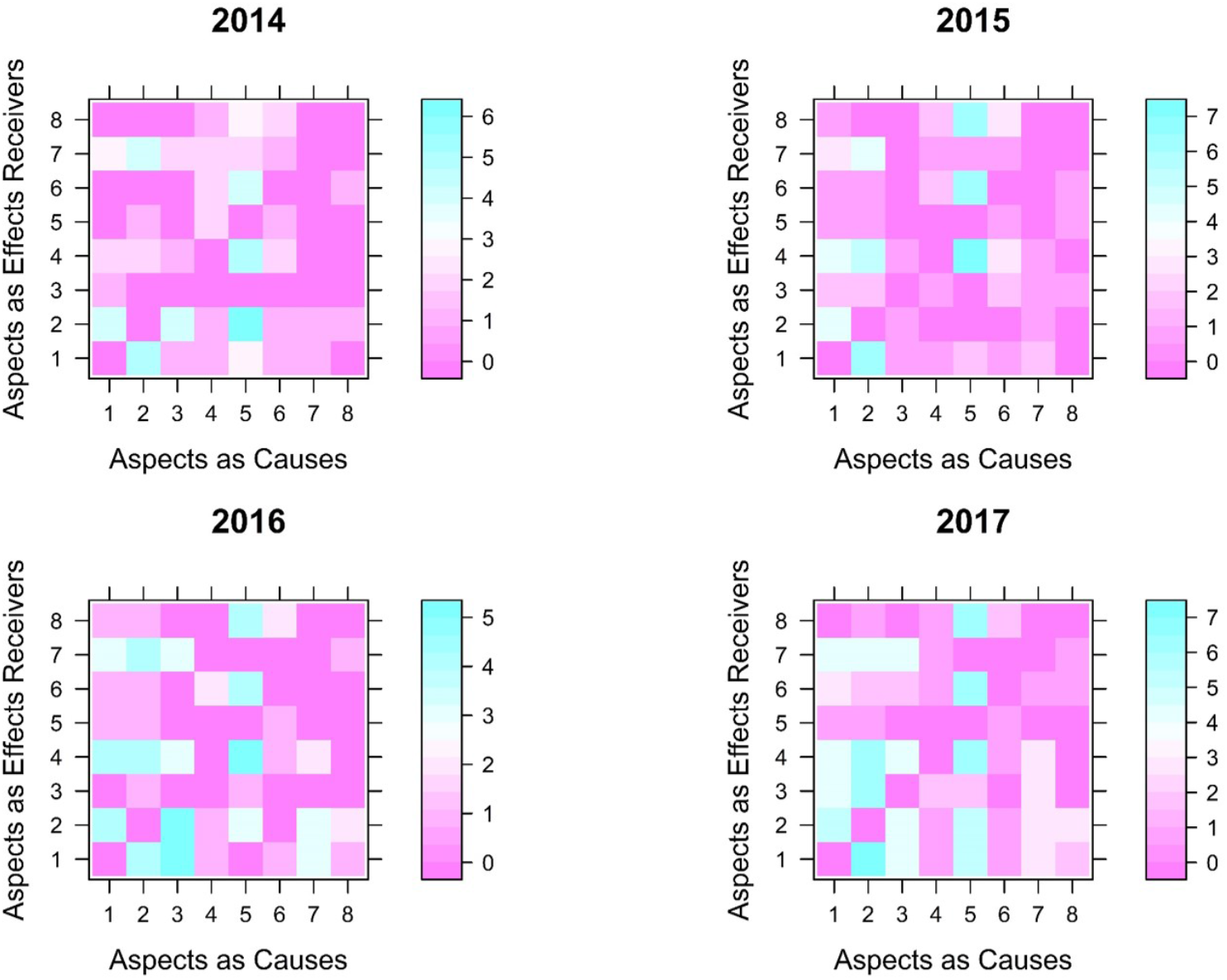
The heatmap of association numbers in W1 from 2014 to 2017

**Figure 3.**
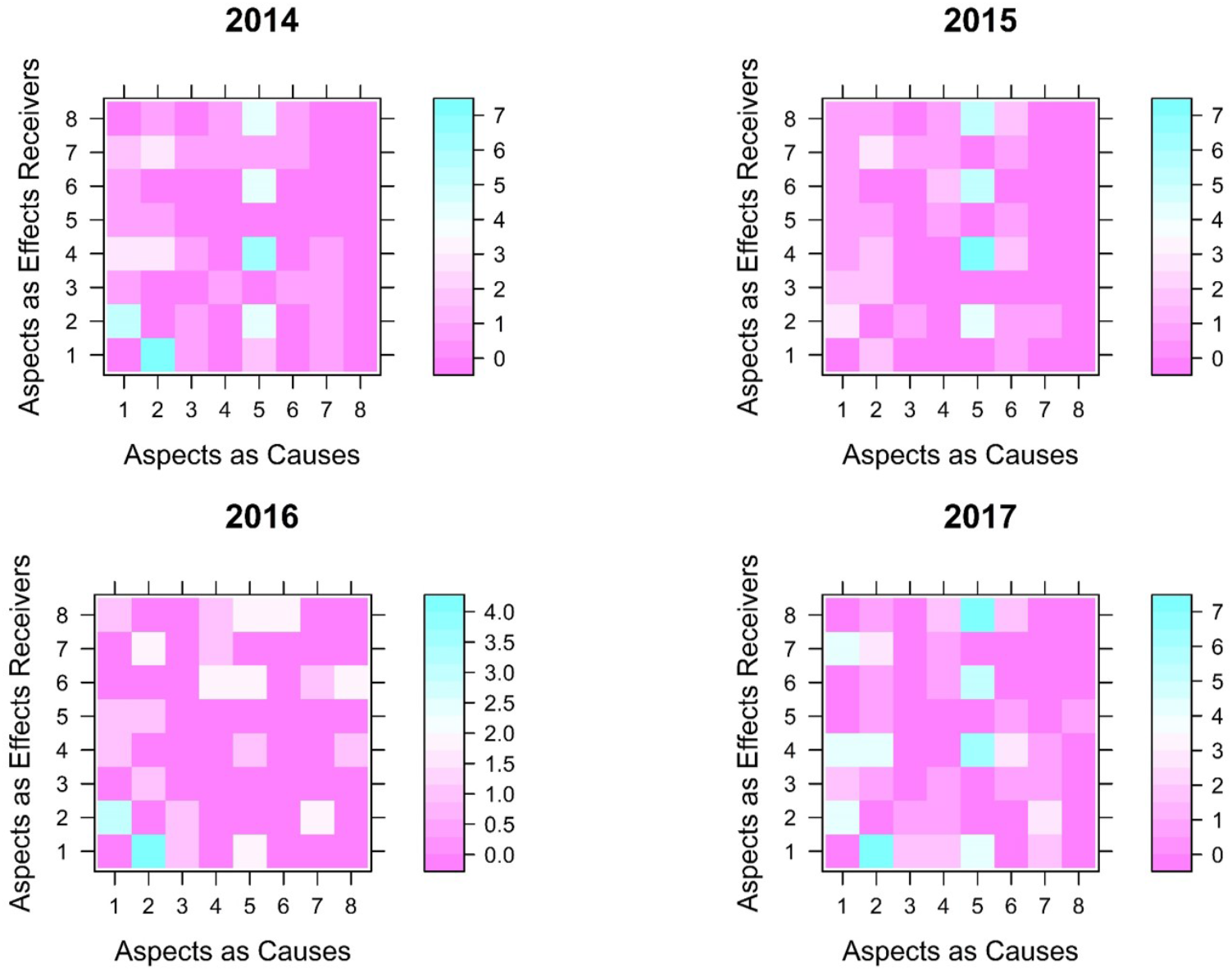
The heatmap of association numbers in W2 from 2014 to 2017

**Figure 4.**
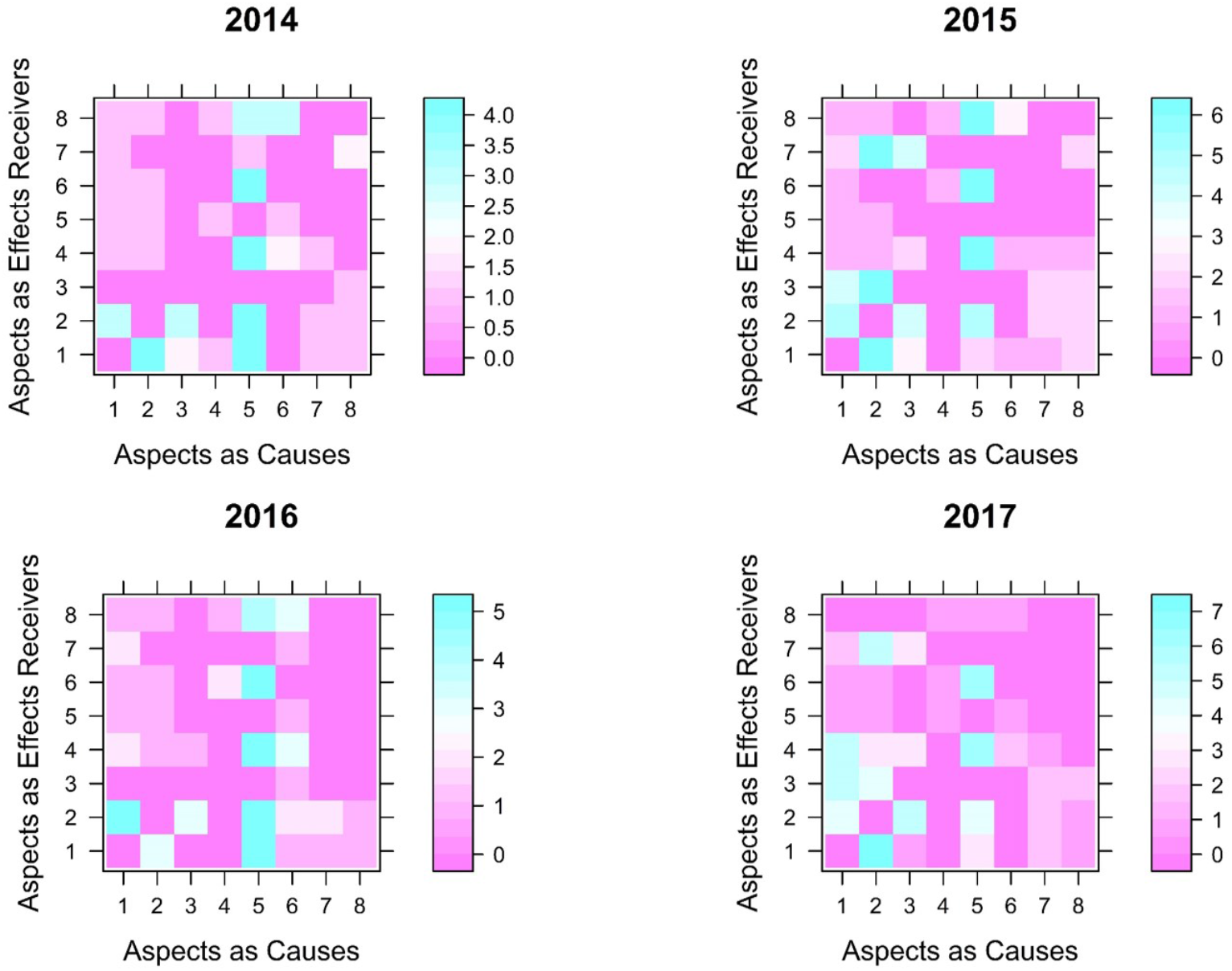
The heatmap of association numbers in W3 from 2014 to 2017

**Figure 5.**
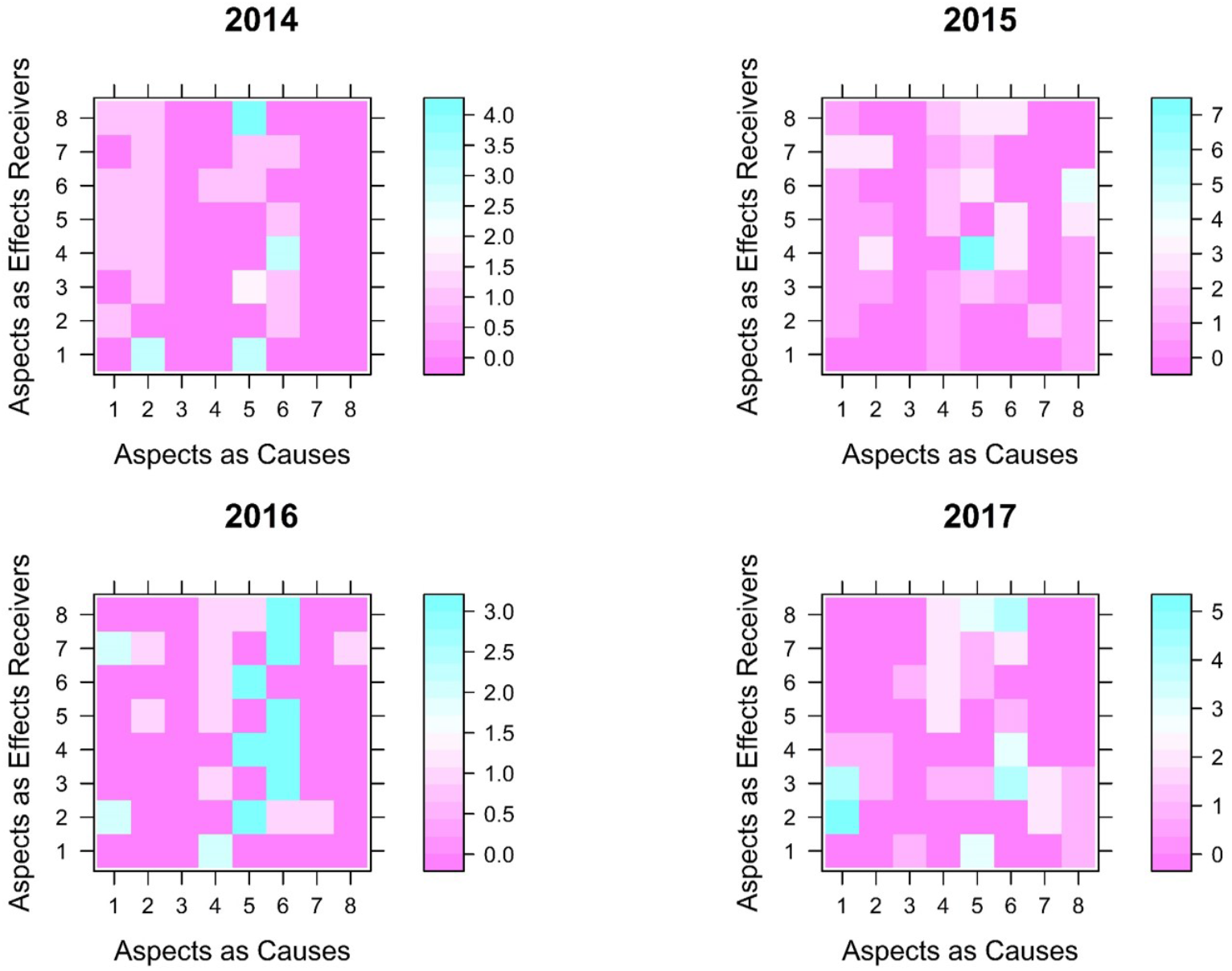
The heatmap of association numbers in W4 from 2014 to 2017

**Figure 6.**
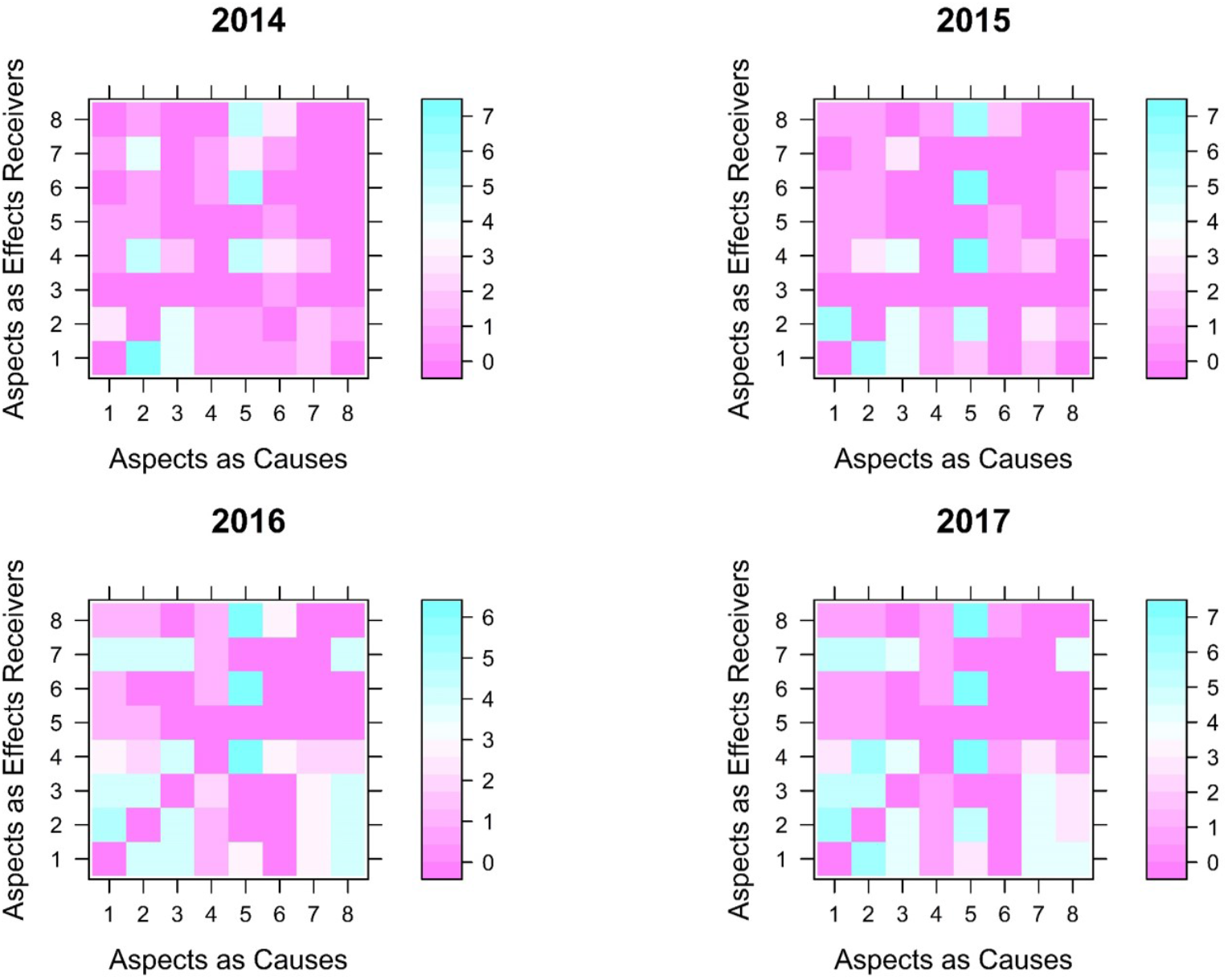
The heatmap of association numbers in W5 from 2014 to 2017

**Figure 7.**
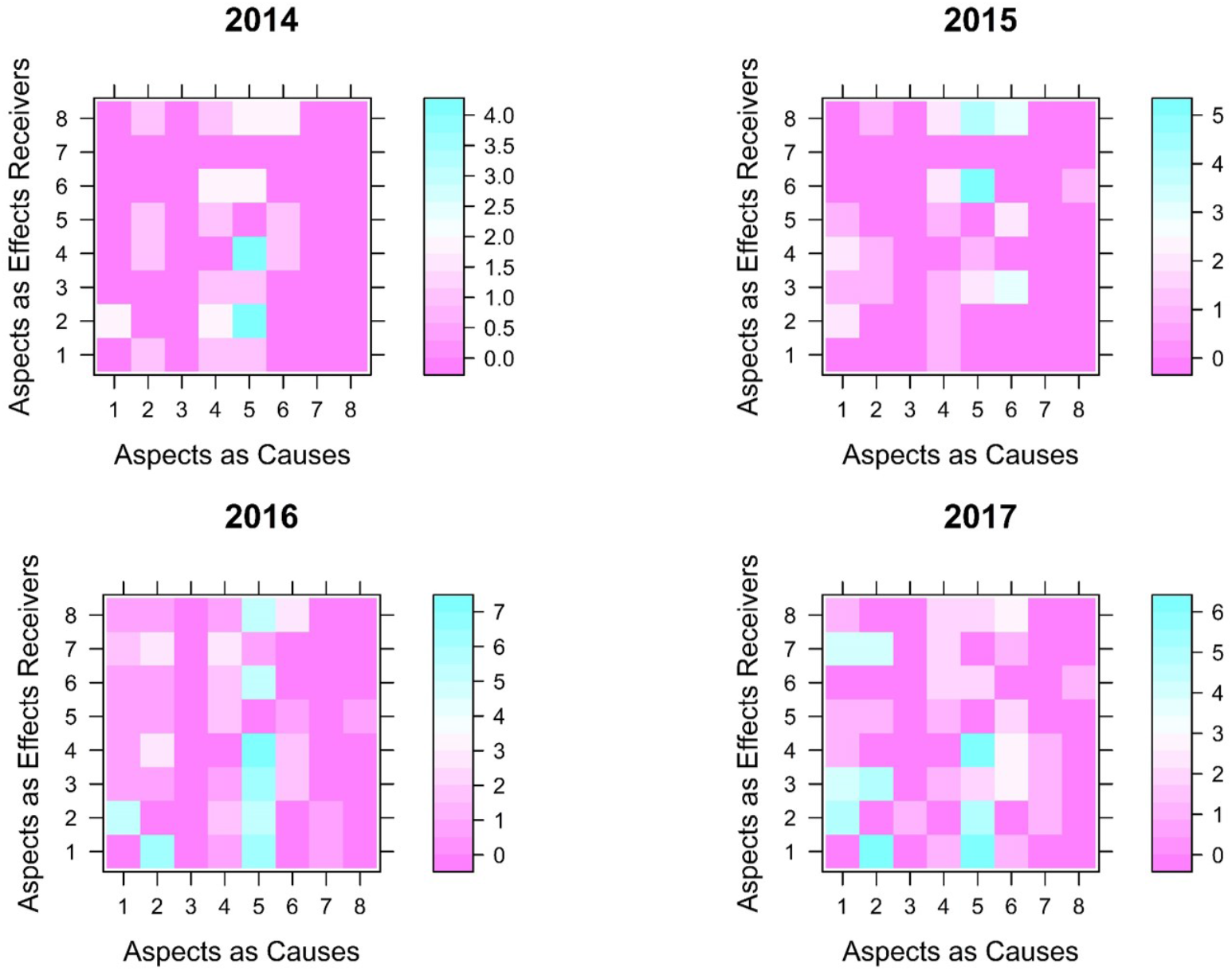
The heatmap of association numbers in W6 from 2014 to 2017

**Figure 8.**
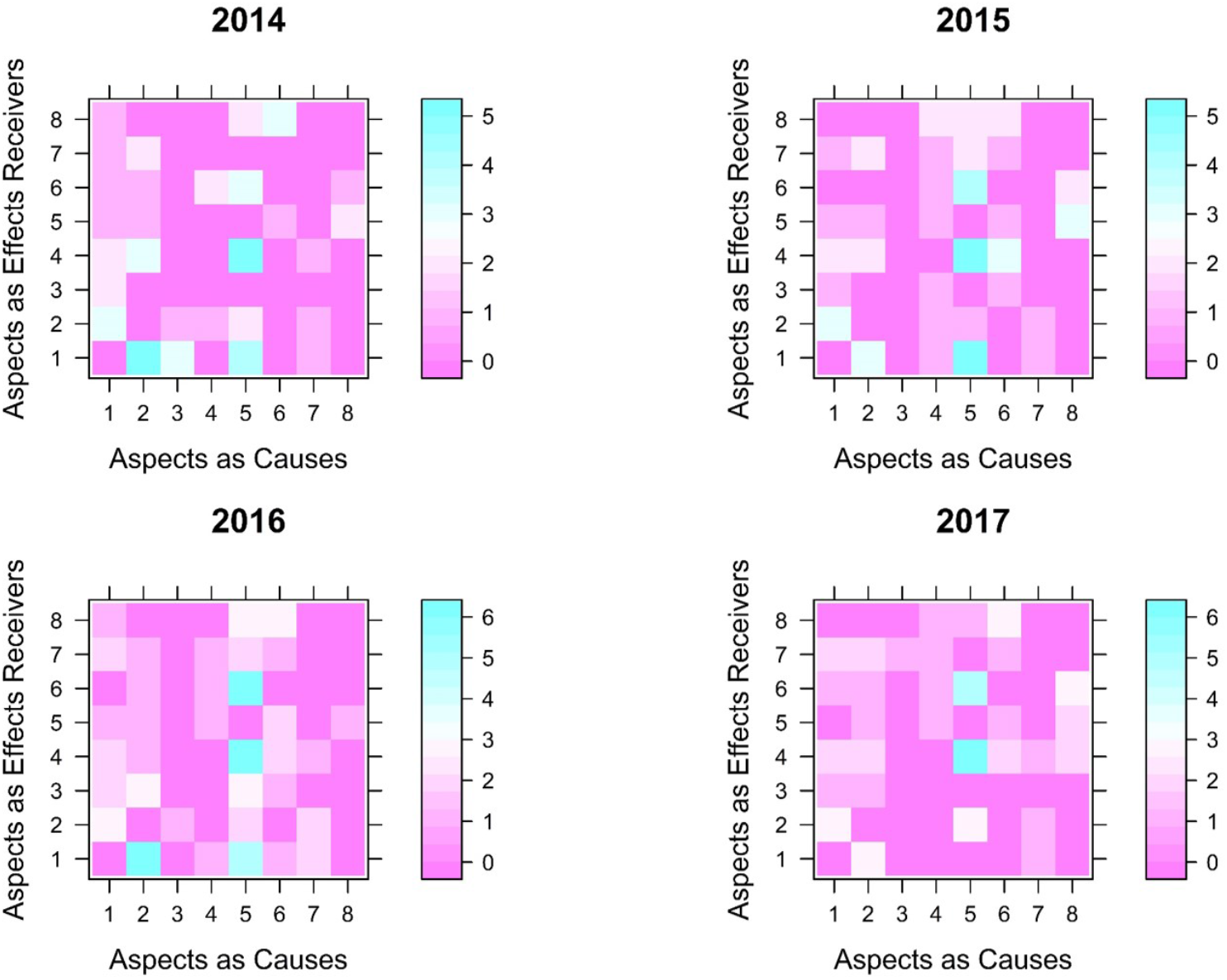
The heatmap of association numbers in W7 from 2014 to 2017

**Figure 9.**
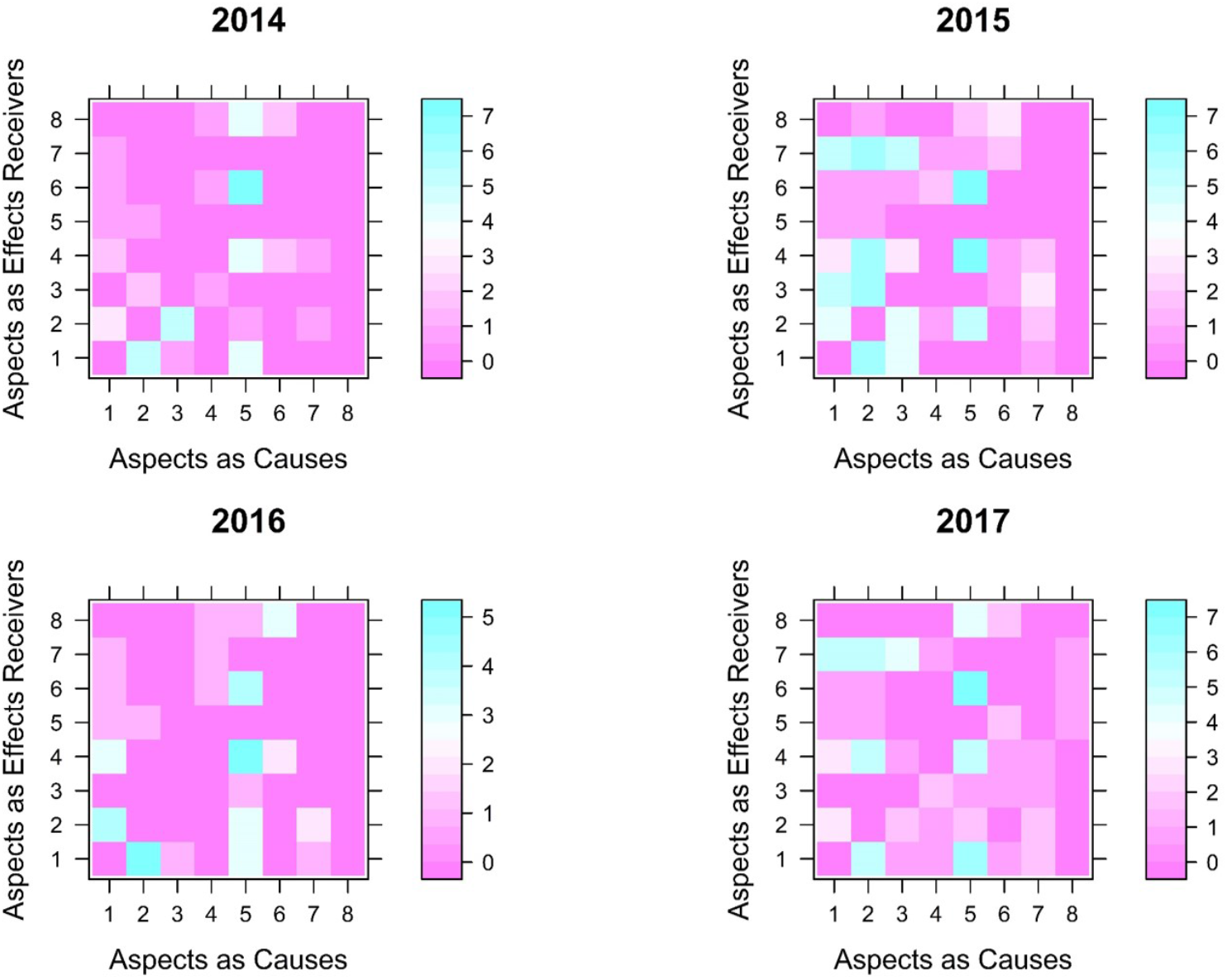
The heatmap of association numbers in W8 from 2014 to 2017

There are some interesting observations associated with invariant status or changing status as desired in the four years for each ward. For instance, aspects as effect receivers are much more possibly to be the most important aspect for a ward in year compared with aspects as causes because 78.8% of the most important aspects for selected wards across four years are effect receivers. Furthermore, associations for wards are not correlated with the performance levels simply since wards with the same performance levels such as W1 and W4 in 2015 are not having same important aspects, roles and associations for all aspects and total number of associations for W1 is decreasing from 2015 to 2016 while its level is changed from B to A in these two years. Several connections exist between trends of levels and trends of some aspects in wards or wards themselves. If the level for a ward is changed from C to B or B to A, the trend of levels is regarded as increasing. If the level for a ward is changed from A to B or B to C, the trend of levels is regarded as decreasing. The trend of levels and trend of total number of associations are the same for W2 and W7. The trend of levels and trend of degree of importance of Safety Climate, Job Satisfaction and Working Conditions are the same for W2. The trend of levels and trend of degree of importance of Working Conditions are the same for W6. The trend of levels and trend of degree of importance of Emotional Exhaustion and Working Conditions are the same for W7. The most or least important aspects are not the same during these four years for each ward. The roles of a few aspects are nearly constant for some selected wards during these four years. Emotional Exhaustion is always an effect receiver for selected wards except W2 and Safety Climate is an effect receiver for W2 and W7 across four years. Stress Recognition is a cause for W1, W3, W5, W7 and W8 and Work and Life Balance is a cause for W1, W2, W6 and W8 across four years. Working Conditions is a cause for W1 and W4 and Job Satisfaction is a cause for W4 and W6 across four years.

## DISCUSSION

There are two potential reasons why the overall evaluated levels are not closely relative with associations with remarkable estimation performance. One possible reason is that the scores for classifying levels present high summation of the information of the numbers of associations. If the degree of importance or role of an aspect is changed and the other aspects in this ward are probably changed accordingly, the performance scores or levels for this ward may remain to be the same as before. Another one is that only the number of associations between aspects is not a crucial factor to decide whether a ward is excellent, good or fair.

### Strengths and limitations of this study

This UB-CF based analysis technique provides a unique manner to understand and analyze probable situations in hospital from SAQ data and SAQ itself. This instrument is contributed to generate a dynamic inner connection with large mean values of accuracies between aspects each year based on different criterions and has a capability to give a suggestion to state important aspects and their roles by recognizing close interrelations between questions and aspects. Therefore, an administration of a hospital may explore some interesting observations from results of this analysis from their perspectives and speculate the appropriate strategies for assisting physicians or nurses in a sub-sector in that hospital to offer services of high quality to patients efficiently.

The two pivotal assumptions of this research are that the data is consistently homogeneous in four years and the aspects are independent to each other. In other words, the SAQ respondents are the same people in these four years and answered with the identical criterion. In addition, safety performance divided into each aspect is affected only by factors pertain to its aspect. These hypothesizes are necessary and beneficial to be applied in advance for this analysis. Nonetheless, they are not possible to exist in this investigation actually, which causes that obvious comparisons between wards do not appear and no distinct summary is extracted from results. Another reason for it is that the communication between researchers and management team in this hospital is absent and relative strategies or messages are not delivered to investigators for conducting a more prominent analysis in this study. Hence, the possible factors causing a phenomenon are difficult to figure out due to its huge potential amount and a safety advancing direction in this hospital is ambiguous.

## CONCLUSION

This research method presents a novel perspective to understand detailed information for specific groups in medical places. Some specific associations between eight aspects for sub-sectors are selected in these years to assess the relationships between performance of sub-sectors and the associations of eight aspects in these sub-sectors. The association analysis in different sub-sectors convey important aspects and their roles for each ward in different years. In addition, there is no convinced evidence to display a strong connection with wards in higher level and features of their distinct associations. The evaluation scores or categories may be determined by the other indicators such as patients’ satisfaction, adverse events, length of hospital day, and readmission rate. One of future research direction is exploring if there exists an accurate and clear relationship between aspects and new evaluated categories in this hospital. Another direction is to integrate staff’s demographic variables (such age, gender, seniority) for further investigating if distinct patterns appear in specific staff groups.

## Data Availability

The data will be available upon approved request.

